# DxToniaNet: A Neural Network for Estimating the Severity of Leg Dystonia in Children with Cerebral Palsy

**DOI:** 10.64898/2026.11.11.26367859

**Authors:** Rishabh Bajpai, Alyssa Rust, Emma Lott, Susie Kim, Sushma Gandham, Keerthana Chintalapati, Joanna Blackburn, Rose Gelineau-Morel, Michael C. Kruer, Dararat Mingbunjerdsuk, Jennifer O’Malley, Laura Tochen, Jeff L. Waugh, Steve Wu, Timothy Feyma, Joel S. Perlmutter, Bhooma R. Aravamuthan

## Abstract

**Importance:** Leg dystonia affects 50–80% of children with cerebral palsy (CP), but severity assessment remains subjective and time-consuming. Objective tools are urgently needed to quantify leg dystonia severity in clinical care and clinical trials.

**Objective:** To develop and validate a deep neural network (DxToniaNet) for leg dystonia severity estimation using 2D clinical video recordings.

**Design, Setting, and Participants:** We retrospectively analyzed video recordings of 298 children with cerebral palsy undergoing a seated hand open-close task as part of routine clinical care (Center 1: 193 videos, 720p/30fps; Center 2: 105 videos, 4K/60fps). Videos were rated for leg dystonia severity by a group of 7 experts using the Global Dystonia Rating Scale (GDRS) leg subscore (range 0-40) which has a minimum clinically important difference (MCID) of 4 points. Videos displaying leg dystonia (median expert GDRS score ≥1) were used to develop and test a machine learning model (DxToniaNet) for estimating leg dystonia severity. Open-source software (OpenPose) estimated bilateral knee, ankle, and toe coordinates, normalized for height and camera positioning, and 69 variance and amplitude features were calculated based on established expert-cited features of leg dystonia. A neural network, DxToniaNet, was developed, trained and validated using 5-fold cross-validation on Center 1 videos before final testing on independent Center 2 videos.

**Main Outcome(s) and Measure(s):** □Mean absolute error (MAE), root mean square error (RMSE), and accuracy in differentiating between videos with clinically meaningful differences in GDRS scores were calculated relative to experts’ median GDRS scores.

**Results:** When estimating experts’ leg GDRS scores in testing data, the proposed machine learning (ML) model yielded a MAE of 1.71 ± 1.68 and RMSE of 2.21 ± 0.34 (significantly less than the GDRS MCID of 4), and 90-94% accuracy in differentiating between videos with greater than a 4-point difference in the GDRS.

**Conclusions and Relevance:** We have demonstrated that DxToniaNet can accurately estimate leg dystonia severity compared to experts’ assessments with a small (clinically insignificant) error margin. Our methodology supports generalizability since DxToniaNet was developed and tested using independent videos datasets from different centers, recorded under different conditions. In the future, DxToniaNet may help facilitate standardized leg dystonia severity estimation in clinical care and clinical trials.

**Key points:** *Question:* Can a deep learning model use clinically-acquired videos to accurately estimate experts’ leg dystonia severity assessments in children with cerebral palsy?

*Findings:* An automated deep learning model (DxToniaNet) trained and tested on clinically-acquired smartphone videos of 298 children with cerebral palsy estimates leg dystonia severity with 90-94% accuracy compared to experts’ assessments.

*Meaning:* DxToniaNet is an objective and free tool to estimate leg dystonia severity and may thus help facilitate standardized tracking of longitudinal treatment responses and clinical trial outcomes.

## Introduction

Dystonia is a debilitating movement disorder that most commonly presents in childhood in association with cerebral palsy (CP).^1–4^ Dystonia is difficult to diagnose, particularly in CP where it often co-exists with spasticity and other motor impairments.^5^ Even after diagnosis, dystonia severity assessment can be difficult. All current assessment tools are qualitative, subjective, and time consuming, limiting their utility during routine clinical care even by expert physicians.^6,7^

Yet, dystonia severity assessment is critical for determining response to treatments. Since between 50-80% of people with CP have some degree of leg dystonia, establishing whether an assessment tool can reliably detect clinically meaningful severity differences is essential for tracking treatment responses. This is particularly valuable for multi-site clinical trials where use of subjective and time-consuming severity scales requires significant investments in training raters and establishing rating consensus across sites. We thus propose establishing a standardized objective leg dystonia severity assessment in people with CP.

We have shown that expert-cited features of leg dystonia during a seated alternating hand open-close task can be quantified in clinically-acquired 2D videos and correlate well with experts’ consensus-based qualitative assessments of leg dystonia severity.^8^ We recently leveraged these findings to develop DxTonia: open-source software that uses a machine learning (ML) model trained on these expert-cited features to detect leg dystonia with 82% accuracy compared to experts’ consensus-based assessment.^9^ We now propose expanding DxTonia to assess leg dystonia severity. We hypothesize that expert-cited leg dystonia features can be used to train deep learning models to estimate leg dystonia severity scores and accurately discriminate between subjects with clinically meaningful differences in leg dystonia severity.

## Methods

### Research protocol approvals and study registrations

The research protocol for this study was reviewed and approved by the Washington University in St. Louis Institutional Review Board on 06/05/2023 (Approval Number: 202102101).

### Participants, movement task, and video acquisition

Video recordings of children with CP performing a seated alternating hand open-close task were collected during routine clinical visits at two centers: Center 1 – Gillette Children’s Hospital (St. Paul, MN, USA), and Center 2 – St. Louis Children’s Hospital (St. Louis, MO, USA). Center 1 recordings were made with a Panasonic AG-AC160AP camera (1920×1080, 30fps) between 01/01/2020 and 12/30/2021. Center 2 recordings were made with a Google Pixel 5 smartphone (3840×2160, 60fps) between 4/20/2023 and 12/14/2023. Inclusion criteria were subjects with clinically-confirmed CP diagnoses who were at least 3 years old to allow for the cognition and attention to attempt the task. Exclusion criteria were subjects unable to attempt the task or not having videos capturing their full body. When two or more videos were available for the same child, only the most recent video was included, so each included video was of a unique subject.

During the alternating hand open-close task, participants sit facing forward with their non-dominant hand resting on their thigh, raise their dominant hand to the level of their face, and then rapidly alternate opening and closing their dominant hand for approximately five seconds. This is then repeated with the other hand. This voluntary upper limb movement can trigger dystonia in the legs, also called leg overflow dystonia as per the 2025 consensus definition.^1^ We have previously shown that the seated alternating hand open-close task is effective for identifying leg dystonia^8,9^ and can, in isolation, be used to estimate leg dystonia severity accurately when compared to severity assessments using a prolonged exam protocol (including lying down, seated upper extremity, seated lower extremity, and gait tasks).^10^

To protect participant privacy, faces in all video recordings were blurred using ShotCut and custom-written open source software (SecurePose).^11^ Only face-blurred videos were used for consensus-based expert video assessment, leg coordinate extraction, and ML model training.

### Consensus-based expert video assessment and its qualitative analysis□

Experts (J.B., R.G.M., M.K., D.M., J.O.M., L.T., J.W., S.W.) were pediatric movement disorders physicians with dystonia expertise from a different US centers with none from Centers 1 or 2. Experts used the Global Dystonia Severity Rating Scale (GDRS)^12^ which we have shown yields high interrater reliability for leg dystonia severity assessment in children and young people with CP^8,13^ and has a minimum clinically important difference (MCID) of 4 points.^14^ The GDRS ranges from 0 (no dystonia) to 10 (severe dystonia) for each body region, with a maximum score of 40 for the legs (upper and lower left and right leg).

Of the 298 videos collected from Centers 1 and 2, 24 from Center 1 and 30 from Center 2 were initially discussed by the expert reviewers across six one-hour Zoom meetings (Zoom Video Communications, Inc.) to ensure rating consensus. This video susbet varied by subjects’ gross motor function impairments and preliminary estimates of leg dystonia severity as determined by a different pediatric movement disorders physician (B.R.A) whose ratings were never revealed to the other reviewers or used for further analysis. Once the experts had determined leg dystonia severity in the initial set of 54 videos together, they independently reviewed the remaining 244 videos for leg dystonia severity. Median leg dystonia severity ratings across all experts were used for further analysis. Since this study is focused on leg dystonia severity and not leg dystonia identification, only videos with leg dystonia (median expert GRDS ≥1) were used for training, validation, and testing of the ML models.

### Leg coordinates extraction and feature selection

We used custom-written software (SecurePose)^11^ incorporating OpenPose^15^ to label and extract the 2D X and Y coordinates for 3 points on each leg in each video frame: midpoint of the patella, midpoint between the medial and lateral malleoli, and the tip of the first toe. This yielded 12 coordinates per frame. Labeling accuracy was verified manually in a subset of the frames (at least 50%). Coordinates were normalized to account for patient height and distance from the camera (using maximum knee-to-ankle distance as a reference) and adjusted to account for camera positioning (i.e. the median pelvis X and Y coordinates for each frame were set to zero).

Leg kinematic features were calculated using these 12 coordinates as per established quantifiable movements used by experts to assess leg dystonia severity:^9,13^ hip flexion/extension, hip adduction/abduction, hip internal/external rotation, knee flexion/extension, ankle dorsiflexion/plantarflexion, and foot inversion/eversion.^9,13^ Because we previously found that the variability and amplitude of leg adduction correlate with expert-assessed leg dystonia severity^8,13^ and can be used to accurately identify the presence or absence of leg dystonia,^9^ we used the variance, minimum, and maximum of all leg kinematic measures (a total of 69 features, Supplementary Table 1) as inputs for training ML models to assess leg dystonia severity.

**Table 1.** The performance (mean absolute error) of CNN-DNN during a grid search.

|  | Without Batch Normalization |  |  | With Batch Normalization |  |  |  |
|---|---|---|---|---|---|---|---|
| Learning Rate Activation Function | 0.01 | 0.001 | 0.0001 | 0.01 | 0.001 | 0.0001 | **Mean MAE (Activation function)** |
| ELU<sup>20</sup> | 1.99±1.88 | 1.64±1.78 | 1.61±1.27 | 1.99±1.36 | 1.63±1.91 | 1.67±1.98 | 1.76±1.70 |
| ReLu<sup>21</sup> | 2.01±1.05 | 1.74±1.95 | 1.63±1.25 | 2±1.44 | 1.78±1.36 | 1.79±1.87 | 1.83±1.49 |
| Sigmoid<sup>22</sup> | 2.11±1.74 | 2.09±1.58 | 2.16±1.66 | 2.14±2.17 | 2.07±1.45 | 2.17±1.25 | 2.12±1.64 |
| LeakyReLU<sup>23</sup> | 1.84±1.65 | **1.54±1.63** | 1.55±1.66 | 1.88±0.54 | 1.56±1.54 | 1.6±1.39 | 1.66±1.55 |
| **Mean MAE (Learning Rate)** | 1.99±1.58 | 1.75±1.74 | 1.74±1.46 | 2.00±1.61 | 1.76±1.57 | 1.81±1.62 |  |
| **Mean MAE (Batch Normalization)** | 1.83±1.59 |  |  | 1.86±1.60 |  |  |  |

### Finding the best performing ML model (DxToniaNet)

We have used conventional ML methods to identifying leg dystonia in people with CP,^9^ but dystonia severity is a more complex and continuous outcome than is simple binary dystonia identification and thus requires modeling complex non-linear relationships. We employed six ML regression algorithms: linear regression, decision tree, SVM, ensemble trees, gaussian process regression, and neural networks. For the neural network algorithm, we evaluated four architectures: (1) dense layers only (DNN), (2) convolutional layers followed by dense layers (CNN⍰DNN), (3) long short⍰term memory layers followed by dense layers (LSTM⍰DNN), and (4) a hybrid CNN–LSTM–DNN model. Model architecture and key hyperparameters (e.g. batch normalization, learning rate, and activation functions) can significantly influence training dynamics and affect model performance. We systematically varied each of these parameters and compared the mean absolute error (MAE) between the model’s estimation and experts’ consensus severity ratings across multiple design alternatives on the validation data (i.e. a grid search).^16,17^ After identification of key hyperparameter values, additional parameters (e.g. number of layers, filter sizes, layer⍰specific options, stride lengths, and pooling window dimensions) were comparably iteratively refined. The best performing regression algorithm, architecture, and hyperparameters were thus identified as the ones that minimized the MAE.

To minimize bias and ensure robust model evaluation, we employed five-fold cross-validation using the scikit-learn^18^ library. Videos from Center 1 were divided into training (2/3), validation (1/6), and testing (1/6) sets, maintaining comparable GDRS score distributions across each set. To account for potential variations due to arbitrary division of the data set, we repeated this process five times, creating five different training, validation, and testing sets. The best performing model across these five different sets was identified as outlined above.

The resultant model (DxToniaNet) was then trained on all Center 1 videos (100 epochs, batch size of 32 samples) using 69 variance and amplitude features and GDRS values standardized with zero mean and one standard deviation. A model checkpoint of early stopping with 20 sample points on validation loss was used to avoid overfitting.

### DxToniaNet generalizability testing

After completing all training, validation, and initial testing exclusively with videos from Center 1, we tested DxToniaNet on an independent external dataset from Center 2. Noting that the GDRS has an MCID of 4 points, we evaluated the model’s accuracy in assessing leg dystonia severity in two ways:

1. Estimating experts’ median GDRS scores: We determined whether the MAE and root mean squared error (RMSE) were significantly less than 4.
2. Comparing DxToniaNet’s estimated GDRS scores in video pairs with clinically meaningful leg dystonia severity differences: In video pairs where experts’ GDRS scores differed by ≥4 points, we determined whether the model correctly assigned the higher GDRS score to the video that experts also rated higher.

### Translation to cloud infrastructure and web application architecture (DxToniaWeb)

To translate the standalone Windows-based DxTonia software and the newly developed DxToniaNet severity estimation model into a clinically accessible tool, we developed DxToniaWeb, a multi-user Flask web application. Unlike the desktop version which requires download and installation on a Windows-based Graphical Processing Unit, the web application allows universal access to a single-click dystonia assessment workflow directly within a web browser interface.

The web application’s processing pipeline incorporates:

1. Video normalization and metadata correction: Uploaded videos undergo automated rotation correction and metadata normalization using FFmpeg and ExifTool, standardizing video orientation across different recording devices.
2. Kinematic feature extraction: Real-time keypoint extraction is executed via a server-side OpenPose queue, yielding leg coordinates (knee, ankle, first toe).
3. Spatiotemporal association and tracking: A motion-based frame-association algorithm tracks the patient across video frames to resolve multi-person clutter.
4. Ensemble classification and inference: Cleaned kinematics are standardized and mapped to the 69 variance and amplitude features, which are then passed to both the original dystonia identification model (DxTonia) and the newly optimized dystonia severity assessment model (DxToniaNet).

To handle these computationally and temporally intensive analysis stages without HTTP timeouts, we implemented a background job queuing architecture orchestrated by Redis and Redis Queue. A dedicated administrator dashboard provides comprehensive operational monitoring, system logging, workflow recovery controls (e.g., job cancellation, re-queuing, and stale state purging), and secure CSV exports of final movement kinematics and dystonia assessments. The RAID 5 encrypted storage, zero-trust security infrastructure, and planned Zitadel integration to ensure robust data protection are detailed in Supplementary eMethods 1 and eMethods 2.

## Results

### Participants and demographics

A total of 193 videos from Center 1 and 105 videos from Center 2 met inclusion and exclusion criteria. After experts analyzed these videos for leg dystonia, 107 videos from Center 1 (55.4%, age 7.1 ± 3.7 years, GDRS range 1.0–21.0) and 58 videos from Center 2 (55.2%, age 10.4 ± 3.8 years, GDRS range 1–14) had leg dystonia and were used for model development (Supplementary Table 2).

### ML algorithm performance comparison for leg dystonia severity estimation

A grid search was performed for each ML algorithm using testing datasets from Center 1 with five-fold cross validation (Supplementary Table 3). The best performing ML algorithm, CNN-DNN, performed best with a learning rate of 0.001, activation function of LeakyReLU, and without batch normalization layers (Table 1). Based on this, the Adam optimizer^19^ with a learning rate of 0.001, epsilon = 10^-7^, and first- and second-moment estimation decay rates of 0.9 and 0.999, respectively, were selected to update the weights of the network during the training.

Averaged across all five folds, the final CNN-DNN model (DxToniaNet) obtained a mean MAE of 1.54±1.63 and RMSE of 2.16±0.33 for the Center 1 validation datasets and a mean MAE of 1.62±1.47 and RMSE of 2.17±0.24 for the Center 1 testing datasets (Table 2). DxToniaNet was then re-trained on the full Center 1 dataset (n=107). Finally, when tested on the independent Center 2 dataset (n=58), DxToniaNet had a mean MAE of 1.71±1.68 and RMSE of 2.21±0.34 (Table 2), both significantly less than the GDRS MCID of 4 (one-sided one-sample t-tests, p< 0.001).

**Table 2.** The performance of machine learning models for estimating GDRS during validation and testing using five different data groupings from Center 1, as well as the performance of the final model (fine-tuned NN on all videos from Center 1) on data from Center 2.

| **Validation performance of ML model from each dataset grouping from Center 1** |  |  |
|---|---|---|
| **Dataset Groupings** | **Validation RMSE** | **Validation MAE** |
| Fold 1 | 3.25 | 1.98 |
| Fold 2 | 1.83 | 1.57 |
| Fold 3 | 1.74 | 1.06 |
| Fold 4 | 1.91 | 1.60 |
| Fold 5 | 2.09 | 1.48 |
| Mean | 2.16±0.62 | 1.54±0.33 |
| **Testing performance of ML model from each dataset grouping from Center 1** |  |  |
| **Dataset Groupings** | **Test RMSE** | **Test MAE** |
| Fold 1 | 1.99 | 1.46 |
| Fold 2 | 2.00 | 1.43 |
| Fold 3 | 1.80 | 1.51 |
| Fold 4 | 2.44 | 1.95 |
| Fold 5 | 2.61 | 1.74 |
| Mean | 2.17±0.34 | 1.62±0.22 |
| **Testing performance of the final fine-tuned model on Center 2** |  |  |
| **Final model** | **Test RMSE** | **Test MAE** |
| All 58 videos | 2.21±0.34 | 1.71±1.68 |

### Comparing DxToniaNet’s estimated GDRS scores in video pairs with clinically meaningful differences in leg dystonia severity

We evaluated how accurately DxToniaNet identifies which of two videos displays more severe leg dystonia when experts’ scores differ by at least the GDRS MCID of 4 points between videos.

This evaluation was conducted across three datasets: the Center 1 validation dataset (which generated 1347 unique video pairs that differed by a GDRS≥4), the Center 1 testing dataset (1467 pairs), and the Center 2 testing dataset (596 pairs). DxToniaNet correctly identified the subject with greater dystonia severity in 1240/1347 pairs (92.1%) from the Center 1 validation dataset, 1315/1467 pairs (89.6%) from the Center 1 testing dataset, and 558/596 pairs (93.6%) from the Center 2 testing dataset.

### Translation Feasibility and System Validation

To validate DxToniaWeb as a robust alternative to local desktop execution (DxToniaNet), we evaluated DxToniaWeb across four domains using both Center 1 and Center 2 datasets:

1. Analytical equivalence: We ran videos from both centers through DxToniaNet and DxToniaWeb with identical results.
2. System performance and computational latency: We measured the execution latency of the Redis and Redis Queue orchestrated pipeline on videos from both centers. We found that the execution latency was 1.2x the video duration for videos recorded at 720p (HD resolution, 30 Hz frame rate), 6.45x the duration for videos recorded at 1080p (full HD resolution, 30 Hz frame rate), and 9.13x the duration for videos recorded at 3840p (4K resolution, 60 Hz frame rate) (Supplementary Table 4).
3. Effect of down-sampling on performance: Running DxToniaWeb on down-sampled videos from Center 2 (720p and 30Hz frame rate) yielded the same MAEs as when using the native videos (3840p and 60Hz frame rate) confirming that down-sampling can reduce execution times without compromising accuracy ) (Supplementary Table 5).
4. Desktop browser compatibility: DxToniaWeb was tested across Google Chrome (version 148), Mozilla Firefox (version 151), Apple Safari (version 18), and Microsoft Edge (version 148) and successfully processed uploaded videos, managed job queuing, and exported dystonia assessments as PDF and CSV files across all platforms.

## Discussion

We present DxToniaNet, a novel CNN-DNN neural network architecture capable of accurately estimating experts’ leg dystonia severity assessments (GDRS scores) with errors significantly less than the threshold for clinically meaningful differences in leg dystonia severity. Videos used for developing the model were completely distinct from videos used for final testing, which were collected at a different medical center under different recording environments and parameters, supporting generalizability. We additionally demonstrate that DxToniaNet can differentiate between videos with clinically meaningful differences in leg dystonia severity with 90-94% accuracy compared to experts’ assessments. Finally, we present DxToniaWeb, a web application that executes the functionalities of DxToniaNet without requiring local installation. Both DxToniaNet and DxToniaWeb were developed and tested using only face-blurred videos, supporting patient privacy if eventually used in clinical settings.

DxToniaNet relies on the integration of experts’ clinical assessments with advanced deep learning. By leveraging expert-cited features (including variance and amplitude of leg adduction as validated in prior work),^8,9,13^ we moved beyond binary dystonia detection to continuous severity estimation. The CNN-DNN architecture (Table 3) effectively captures spatial hierarchies in movement features, outperforming conventional ML methods (Supplementary Table 3). The grid search optimization (Table 1) confirmed that LeakyReLU activation and Adam optimization with a learning rate of 0.001 were critical for minimizing error, while batch normalization provided only marginal benefit.

**Table 3.** Architecture of the final ML model.

| **S.No.** | **Layer Type** | **Activation function** | **Filter size** | **Layer parameter** | **Stride** | **Output size** | **Parameters** |
|---|---|---|---|---|---|---|---|
| 1 | Convolution 1 | LeakyReLU | 9X1 | 32 filters | 1 | 61X32 | 320 |
| 2 | Convolution 2 | LeakyReLU | 7X1 | 128 filters | 1 | 55X128 | 28800 |
| 3 | Convolution 3 | LeakyReLU | 5X1 | 64 filters | 1 | 51X64 | 41024 |
| 4 | Convolution 4 | LeakyReLU | 3X1 | 64 filters | 1 | 49X64 | 12352 |
| 5 | Flatten |  |  |  |  | 3136 |  |
| 6 | Dense 1 | LeakyReLU |  | 16 neurons |  | 16 | 50192 |
| 7 | Dense 2 | LeakyReLU |  | 32 neurons |  | 32 | 544 |
| 8 | Dense 3 | LeakyReLU |  | 128 neurons |  | 128 | 4224 |
| 9 | Dense 4 | Linear |  | 128 neurons |  | 128 | 129 |

The development of DxToniaWeb facilitates the potential translation of deep learning models into clinical practice. While the original desktop version of DxTonia provided an open-source solution for automated dystonia identification, it was constrained by platform dependencies (Windows-only execution) and localized computational requirements (requiring dedicated GPU hardware), which limit its use in typical clinical environments. By integrating both the DxTonia leg dystonia identification model and the new DxToniaNet leg dystonia severity assessment model into a secure, browser-accessible Flask pipeline, DxToniaWeb democratizes access to quantitative movement analysis. The architecture’s RAID 5 encrypted storage, zero-trust infrastructure, TSL for data transfer, and planned Zitadel integration ensure robust data protection.

### Limitations and future directions

DxToniaNet relies on videos of the seated hand open-close task to isolate overflow leg dystonia, which may not capture all dystonia presentations and excludes children unable to perform the task. Performance variability across Centers 1 and 2 (with differing video resolutions, durations, and demographics) underscores the model’s sensitivity to data characteristics, though robust cross-center validation mitigates this concern. Future work must increase generalizability by expanding validation to other clinically relevant tasks (e.g., gait) and other populations with greater gross motor impairments or with dystonia due to other etiologies. Future validation should also include longitudinal evaluation of the same subject across videos (e.g. pre- and post-treatment to assess responsiveness). This validation could support use of DxToniaNet to facilitate standardized, automated, and accurate leg dystonia assessment in multi-site clinical trials.

### Conclusions

DxToniaNet is a new CNN-DNN model that estimates leg dystonia severity scores using only face-blurred videos with an error less than the clinically meaningful threshold. It was trained and tested on distinct video data sets from two different medical centers, supporting generalizability, and it can distinguish clinically meaningful leg dystonia severity differences with 90-94% accuracy. DxToniaWeb is a secure web-based implementation of DxToniaNet which does not require local download or specialized hardware, thus facilitating broad use. Future studies will focus on expanding DxToniaNet and DxToniaWeb to assess other tasks and assess responsiveness to dystonia treatments.

## Data Availability

All data produced in the present study are available upon reasonable request to the authors.

## Acknowledgment

This project is supported by Pediatric Epilepsy Research Foundation Registry Infrastructure Grant and the National Institute of Neurological Disorders and Stroke (NINDS 1K08NS117850-01A1 and NINDS 5R01NS140701).

## References

1. Albanese, A. et al. Definition and Classification of Dystonia. Mov. Disord. 40, 1248–1259 (2025).

2. McIntyre, S. et al. Global prevalence of cerebral palsy: A systematic analysis. Dev. Med. Child Neurol. 64, 1494–1506 (2022).

3. Rice, J., Skuza, P., Baker, F., … R. R.-… M. & C. & 2017, undefined. Identification and measurement of dystonia in cerebral palsy. Wiley Online Libr. Rice, P Skuza, F Bak. R Russo, D FehlingsDevelopmental Med. Child Neurol. 2017•Wiley Online Libr. 59, 1249–1255 (2017).

4. Steeves, T. D., Day, L., Dykeman, J., Jette, N. & Pringsheim, T. The prevalence of primary dystonia: A systematic review and meta-analysis. Mov. Disord. 27, 1789–1796 (2012).

5. Aravamuthan, B., Landsness, E. C. & Silbermann, E. ANA Webinars: implementation of a conference-based virtual networking event. Ann. Clin. Transl. Neurol. 8, 525–528 (2021).

6. Aravamuthan, B., Pearson, T. S., Chintalapati, K. & Ueda, K. Under-recognition of leg dystonia in people with cerebral palsy. Ann. Child Neurol. Soc. 1, 162–167 (2023).

7. Aravamuthan, B. R. et al. Multicenter Improvement in Screening for Dystonia in Young People With Cerebral Palsy. Neurol. Clin. Pract. 15, e200469 (2025).

8. Gemperli, K. et al. Chronic Striatal Cholinergic Interneuron Excitation Causes Cerebral Palsy-Related Dystonic Behavior in Mice. Ann. Neurol. (2025) doi:10.1002/ANA.27299.

9. Bajpai, R. et al. Using Expert-Cited Features to Detect Leg Dystonia in Cerebral Palsy. Ann. Neurol. (2026) doi:10.1002/ana.78130.

10. Lott, E. et al. Tasks for assessing dystonia in young people with cerebral palsy. Ped Neurol 2025.09.07.25334597 at 10.1101/2025.09.07.25334597 (2025).

11. Bajpai, R. & Aravamuthan, B. SecurePose: Automated Face Blurring and Human Movement Kinematics Extraction from Videos Recorded in Clinical Settings. arXiv (2025) doi:10.48550/arXiv.2402.14143.

12. Comella, C. L., Leurgans, S., Wuu, J., Glenn, S. T. & Chmura, T. Rating scales for dystonia: a multicenter assessment. Mov. Disord. 18, 303–312 (2003).

13. Aravamuthan, B. R. et al. Determinants of gait dystonia severity in cerebral palsy. Dev. Med. Child Neurol. 65, 1–10 (2023).

14. Lott, E. et al. The Relationship Between Limb Dystonia Severity and Functional Impact in Children With Cerebral Palsy. J. Child Neurol. (2026) doi:10.1177/08830738261477966;WGROUP:STRING:PUBLICATION.

15. Cao, Z., Hidalgo, G., Simon, T., Wei, S. E. & Sheikh, Y. OpenPose: Realtime Multi-Person 2D Pose Estimation using Part Affinity Fields. IEEE Trans. Pattern Anal. Mach. Intell. 43, 172–186 (2018).

16. Bajpai, R. & Joshi, D. Foot2hip: A Deep Neural Network Model for Predicting Lower Limb Kinematics From Foot Measurements. IEEE/ASME Trans. Mechatronics 28, 2248–2258 (2023).

17. Bajpai, R., Khare, S. & Joshi, D. A Multimodal Model-Fusion Approach for Improved Prediction of Freezing of Gait in Parkinson’s Disease. IEEE Sens. J. 23, 16168–16175 (2023).

18. Pedregosa, F. et al. Scikit-learn: Machine Learning in Python. J. Mach. Learn. Res. 12, 2825–2830 (2012).

19. Kingma, D. P. & Ba, J. L. Adam: A Method for Stochastic Optimization. 3rd Int. Conf. Learn. Represent. ICLR 2015 - Conf. Track Proc. (2014).

20. Clevert, D. A., Unterthiner, T. & Hochreiter, S. Fast and Accurate Deep Network Learning by Exponential Linear Units (ELUs). 4th Int. Conf. Learn. Represent. ICLR 2016 - Conf. Track Proc. (2015).

21. Nair, V. & Hinton, G. E. Rectified linear units improve Restricted Boltzmann machines. in ICML 2010 - Proceedings, 27th International Conference on Machine Learning (2010).

22. Narayan, S. The generalized sigmoid activation function: Competitive supervised learning. Inf. Sci. (Ny). 99, 69–82 (1997).

23. Maas, A. L., Hannun, A. Y. & Ng, A. Y. Rectifier Nonlinearities Improve Neural Network Acoustic Models. (2013).

