## Supplement Table 1 for "DxToniaNet: A Neural Network for Estimating the Severity of Leg Dystonia in Children with Cerebral Palsy"

### Supplementary Table 1. List of sixty-nine features that were used train ML models. |x| represents absolute value of x.

| Feature index | Features | Feature index | Features |
| --- | --- | --- | --- |
| 1 | Variance Right Toe (Y) | 36 | Minimum \| Right Toe (Y) – Right Ankle (Y) \| |
| 2 | Variance Right Toe (X) | 37 | Minimum \| Right Toe (X) – Right Knee (X) \| |
| 3 | Variance Right Knee (Y) | 38 | Minimum \| Right Toe (X) – Right Ankle (X) \| |
| 4 | Variance Right Knee (X) | 39 | Minimum \| Left Toe (Y) – Left Knee (Y) \| |
| 5 | Variance Right Ankle (Y) | 40 | Minimum \| Left Toe (Y) – Left Ankle (Y)\| |
| 6 | Variance Right Ankle (X) | 41 | Minimum \| Left Toe (X) – Right Toe (X) \| |
| 7 | Variance Left Toe (Y) | 42 | Minimum \| Left Toe (X) – Left Knee (X) \| |
| 8 | Variance Left Toe (X) | 43 | Minimum \| Left Toe (X) – Left Ankle (X) \| |
| 9 | Variance Left Knee (Y) | 44 | Minimum \| Left Knee (X) – Right Knee (X) \| |
| 10 | Variance Left Knee (X) | 45 | Minimum \| Left Ankle (X) – Right Ankle (X) \| |
| 11 | Variance Left Ankle (Y) | 46 | Minimum Left Toe (Y) |
| 12 | Variance Left Ankle (X) | 47 | Maximum Right Toe (Y) |
| 13 | Variance \|Right Toe (X) – Right Ankle (X)\| | 48 | Maximum Right Toe (X) |
| 14 | Variance \| Right Toe (Y) – Right Knee (Y) \| | 49 | Maximum Right Knee (Y) |
| 15 | Variance \| Right Toe (Y) – Right Ankle (Y) \| | 50 | Maximum Right Knee (X) |
| 16 | Variance \| Right Toe (X) – Right Knee (X) \| | 51 | Maximum Right Ankle (Y) |
| 17 | Variance \| Left Toe (Y) – Left Knee (Y) \| | 52 | Maximum Right Ankle (X) |
| 18 | Variance \| Left Toe (Y) – Left Ankle (Y) \| | 53 | Maximum Left Toe (Y) |
| 19 | Variance \| Left Toe (X) – Right Toe (X) \| | 54 | Maximum Left Toe (X) |
| 20 | Variance \| Left Toe (X) - Left Knee (X) \| | 55 | Maximum Left Knee (Y) |
| 21 | Variance \| Left Toe (X) – Left Ankle (X) \| | 56 | Maximum Left Knee (X) |
| 22 | Variance \| Left Knee (X) – Right Knee (X) \| | 57 | Maximum Left Ankle (Y) |
| 23 | Variance \| Left Ankle (X) – Right Ankle (X) \| | 58 | Maximum Left Ankle (X) |
| 24 | Minimum Right Toe (Y) | 59 | Maximum \| Right Toe (Y) – Right Knee (Y) \| |
| 25 | Minimum Right Toe (X) | 60 | Maximum \| Right Toe (Y) – Right Ankle (Y) \| |
| 26 | Minimum Right Knee (Y) | 61 | Maximum \| Right Toe (X) – Right Knee (X) \| |
| 27 | Minimum Right Knee (X) | 62 | Maximum \| Right Toe (X) – Right Ankle (X) \| |
| 28 | Minimum Right Ankle (Y) | 63 | Maximum \| Left Toe (Y) – Left Knee (Y) \| |
| 29 | Minimum Right Ankle (X) | 64 | Maximum \| Left Toe (Y) – Left Ankle (Y) \| |
| 30 | Minimum Left Toe (X) | 65 | Maximum \| Left Toe (X) – Right Toe (X) \| |
| 31 | Minimum Left Knee (Y) | 66 | Maximum \| Left Toe (X) – Left Knee (X) \| |
| 32 | Minimum Left Knee (X) | 67 | Maximum \| Left Toe (X) – Left Ankle (X) \| |
| 33 | Minimum Left Ankle (Y) | 68 | Maximum \| Left Knee (X) – Right Knee (X) \| |
| 34 | Minimum Left Ankle (X) | 69 | Maximum \| Left Ankle (X) – Right Ankle (X) \| |
| 35 | Minimum \| Right Toe (Y) – Right Knee (Y) \| |  |  |
