## Supplement Table 2 for "DxToniaNet: A Neural Network for Estimating the Severity of Leg Dystonia in Children with Cerebral Palsy"

### Supplementary eTable 2. Demographic characteristics of children with CP in videos assessed for leg dystonia.

|  | Center 1 | | | | Center 2 | | | |
| --- | --- | --- | --- | --- | --- | --- | --- | --- |
|  | No dystonia (GDRS < 1) N = 86 | GDRS 1–<4 N = 54 | GDRS ≥ 4 N = 53 | Missing Data (n) | No dystonia (Legs GDRS < 1) N = 47 | Legs GDRS 1–<4 N = 29 | Legs GDRS ≥ 4 N = 29 | Missing Data (n) |
| Age (mean±SD) | 7.5±3.3 | 7.4±4.4 | 6.9±2.9 | 4 | 11.6±3.6 | 10.5±3.9 | 10.2±3.7 | 0 |
| Gestational (mean±SD) | 32.0±6.4 | 31.4±5.7 | 32.2±5.6 | 9 | 33.4±6.6 | 34.9±6.1 | 35.6±5.4 | 9 |
| CP distribution (n, %) |  |  |  | 4 |  |  |  | 1 |
| Hemiplegia | 7, 8% | 3, 6% | 4, 8% |  | 14, 30% | 14, 48% | 16, 57% |  |
| Diplegia | 57, 68% | 29, 56% | 25, 47% |  | 21, 45% | 5, 17% | 7, 25% |  |
| Triplegia | 8, 10% | 14, 27% | 8, 15% |  | 2, 4% | 2, 7% | 3, 11% |  |
| Quadriplegia | 3, 4% | 2, 4% | 14, 26% |  | 8, 17% | 6, 21% | 2, 7% |  |
| Other / none | 9, 11% | 4, 8% | 2, 4% |  | 2, 4% | 2, 7% | 0, 0% |  |
| GMFCS (n, %) |  |  |  | 18 |  |  |  | 0 |
| I | 16, 22% | 14, 27% | 5, 10% |  | 20, 43% | 14, 48% | 10, 34% |  |
| II | 35, 48% | 16, 31% | 21, 41% |  | 23, 49% | 15, 52% | 16, 55% |  |
| III | 18, 25% | 18, 35% | 17, 33% |  | 4, 9% | 0, 0% | 3, 10% |  |
| IV | 4, 5% | 3, 6% | 8, 16% |  | 0, 0% | 0, 0% | 0, 0% |  |
| V | 0, 0% | 0, 0% | 0, 0% |  | 0, 0% | 0, 0% | 0, 0% |  |
| Leg spasticity (n, %) |  |  |  | 12 |  |  |  | 0 |
| Spasticity absent | 6, 8% | 2, 4% | 6, 12% |  | 8, 17% | 4, 14% | 2, 7% |  |
| Spasticity present | 73, 92% | 48, 96% | 46, 88% |  | 39, 83% | 25, 86% | 27, 93% |  |
| Video Duration | 7.2 ± 3.5 seconds | | | | 58.5 ± 5.8 seconds | | | |
| GDRS – Global Dystonia Rating Scale (Center 1: total score; Center 2: legs score). GMFCS – Gross Motor Function Classification System (I–II independently ambulatory without assistive devices, III independently ambulatory with assistive devices, IV–V predominantly uses wheelchair for mobility). Percentages use non-missing cases in each group. | | | | | | | | |
