## Supplement Table 3 for "DxToniaNet: A Neural Network for Estimating the Severity of Leg Dystonia in Children with Cerebral Palsy"

### Supplementary eTable 3. Properties and performance of the ML tested for leg dystonia severity estimation on the testing dataset of the Center 1 using five-fold cross-validation.

| **Algorithm name** | **Model type** | **Hyperparameters*** | **Mean test MAE (GDRS)** |
| --- | --- | --- | --- |
| Linear Regression | Linear | - | 2.5 |
|  | Interaction Linear | - | 5.1 |
|  | Robust Linear | - | 2.4 |
| Decision Tree | Fine Tree | Minimum leaf size = 4 | 2.7 |
|  | Medium Tree | Minimum leaf size = 12 | 2.3 |
|  | Coarse Tree | Minimum leaf size = 36 | 2.3 |
| Support Vector Machine (SVM) | Linear SVM | Kernel = Linear | 2.2 |
|  | Quadratic SVM | Kernel = Quadratic | 3.0 |
|  | Cubic SVM | Kernel = Cubic | 5.0 |
|  | Gaussian SVM | Kernel = Gaussian | 2.3 |
|  | Medium Gaussian SVM | Kernel = Gaussian, Kernel scale = 8.3 | 2.0 |
|  | Coarse Gaussian SVM | Kernel = Gaussian, Kernel scale = 33 | 2.1 |
| Ensemble Trees | Boosted Trees | Minimum leaf size = 8, number of learners = 30, LR = 0.1 | 2.1 |
|  | Bagged Trees | Minimum leaf size = 8, number of learners = 30 | 2.1 |
| Gaussian Process Regression (GPR) | Squared Exponential GPR | Basis function = constant | 2.0 |
|  | Matern 5/2 GPR | Basis function = constant | 2.0 |
|  | Exponential | Basis function = constant | 1.9 |
|  | Rational Quadratic GPR | Basis function = constant | 2.0 |
| Neural Network | DNN | Number of DNN layers = 5 | 5.4 |
|  | CNN-DNN | Number of CNN layers = 4, number of DNN layers = 4 | **1.5** |
|  | LSTM-DNN | Number of LSTM layers = 2, number of DNN layers = 3 | 6.1 |
|  | CNN-LSTM-DNN | Number of CNN layers = 4, number of LSTM layers = 1, number of DNN layers = 5 | 2.4 |
| Note: * Only hyperparameters of the top performing models in each category are shown in the table. Each algorithm was validated during 5 fold cross-validation to find the best-performing hyperparameters. | | | |
