## Supplement Table 4 for "DxToniaNet: A Neural Network for Estimating the Severity of Leg Dystonia in Children with Cerebral Palsy"

### Supplementary eTable 4. Pipeline stage processing latency breakdown (s)

| Video source | Video rotation (FFmpeg) | OpenPose (GPU) | Tracking & patient detection | Dystonia detection ensemble + DxToniaNet inference | Total processing | Real-Time Factor (Processing time/ duration) |
| --- | --- | --- | --- | --- | --- | --- |
| Center 1 (1080p @ 30fps) | 1.14±0.50 | 19.94±8.78 | 2.56±1.13 | 22.80±10.03 | 46.45±20.44 | 6.45±0.01 |
| Center 2 (4K @ 60fps) | 84.6±8.04 | 414.54±39.38 | 9.87±0.94 | 25.38±2.41 | 534.39±50.77 | 9.14±0.02 |
| Center 2 (Down sampled to 720p @ 30fps) | 8.46±0.80 | 36.66±3.48 | 4.23±0.40 | 21.15±2.00 | 70.50±6.70 | 1.21±0.01 |
