## Supplement Table 5 for "DxToniaNet: A Neural Network for Estimating the Severity of Leg Dystonia in Children with Cerebral Palsy"

### Supplementary eTable 5. Effect on DxToniaNet on video down scaling

| Downscaling Target | Mean Processing Latency (s) | Mean Absolute Error (MAE) |
| --- | --- | --- |
| Native 4K (3840x2160 @ 60fps) | 534.39±50.77 | 1.71±1.68 (Reference) |
| Full HD (1920x1080 @ 30fps) | 70.50±6.70 | 1.71±1.68 |
