## Supplement Material for "DxToniaNet: A Neural Network for Estimating the Severity of Leg Dystonia in Children with Cerebral Palsy"

### Supplementary Material

**eMethods 1.1: DxTonia web storage architecture and backup system**

The DxTonia Web application employs a robust storage architecture designed for reliability and data protection. The primary storage utilizes a RAID 5 configuration comprising four 24TB disks, providing 72TB of usable storage with fault tolerance against a single drive failure. All data is stored with comprehensive AES-256 encryption at the filesystem level, ensuring that even direct physical access to the storage media renders the data unreadable without the appropriate decryption keys. The backup system implements an incremental blob-based strategy, where only modified data blocks are synchronized to the backup repository, significantly reducing storage overhead and transfer times. Backups are retained for 30 days with automated expiration, after which all user-identifiable information is systematically purged from both active storage and backup archives.

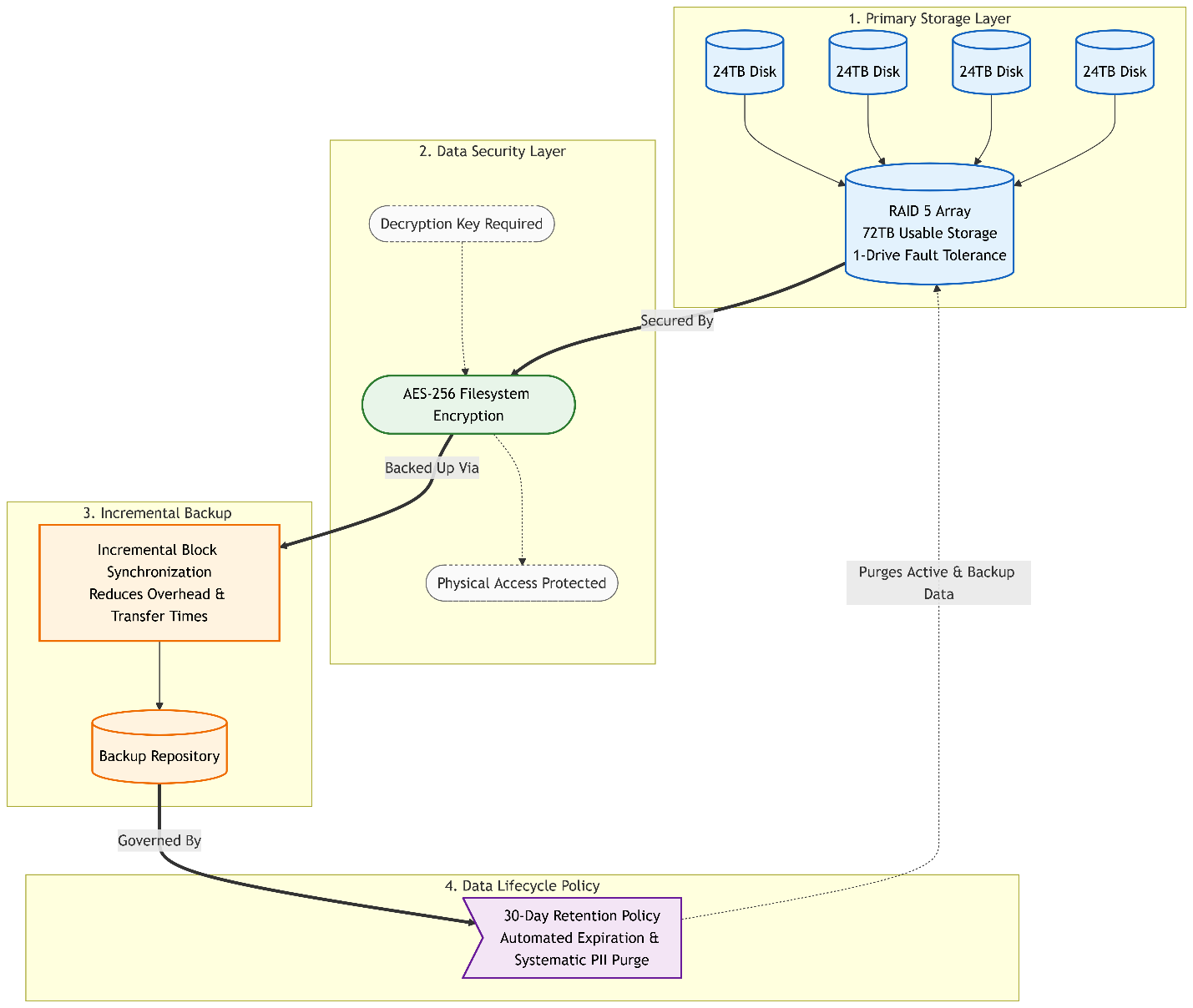

**eMethods 1.2: Security and isolation**

The application operates within Docker containers with strict network isolation, exposing only essential mounted volumes and explicitly permitted network ports. This defense-in-depth approach ensures that the application has no default access to the broader network and can only communicate through controlled channels. All user data, including uploaded videos, processed results, and system logs, is encrypted at rest using AES-256 encryption. The database contains no unencrypted personally identifiable information; all user email addresses, session tokens, and metadata are stored with field-level encryption. The deployment architecture follows the principle of least privilege, with granular access controls preventing unauthorized data access even in the event of container compromise.

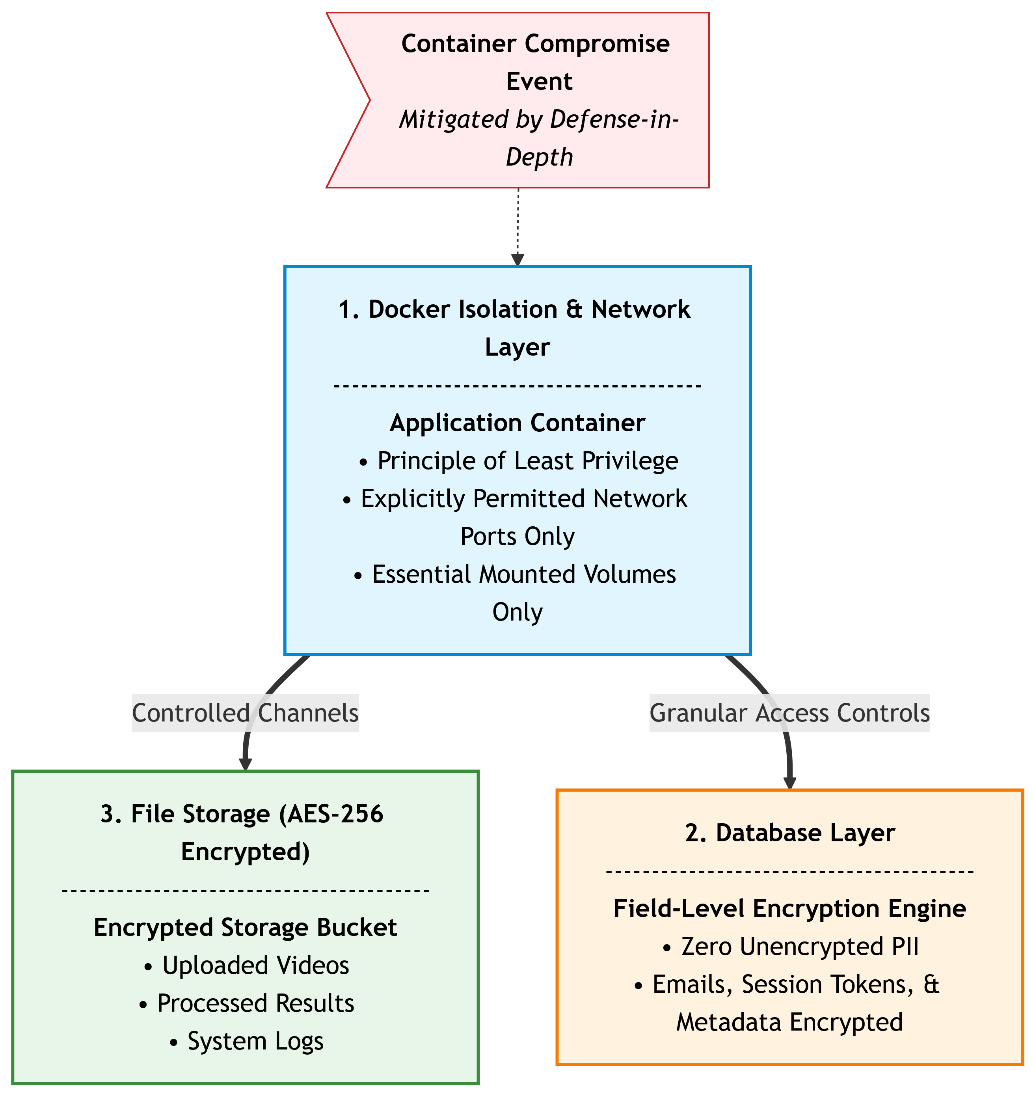

**eMethods 1.3: User data management and privacy**

Users maintain complete control over their data with the ability to request complete deletion of all associated information from the system. The application supports user-initiated data removal, which purges uploads, processed artifacts, analysis results, and backup copies from all storage tiers. The 30-day backup retention policy ensures that user information naturally expires from the system over time, supporting compliance with data minimization principles. Project isolation ensures that users can only access their own data, with per-user directory structures keeping uploads, processed videos, intermediate artifacts, and final results completely separated at the filesystem level.

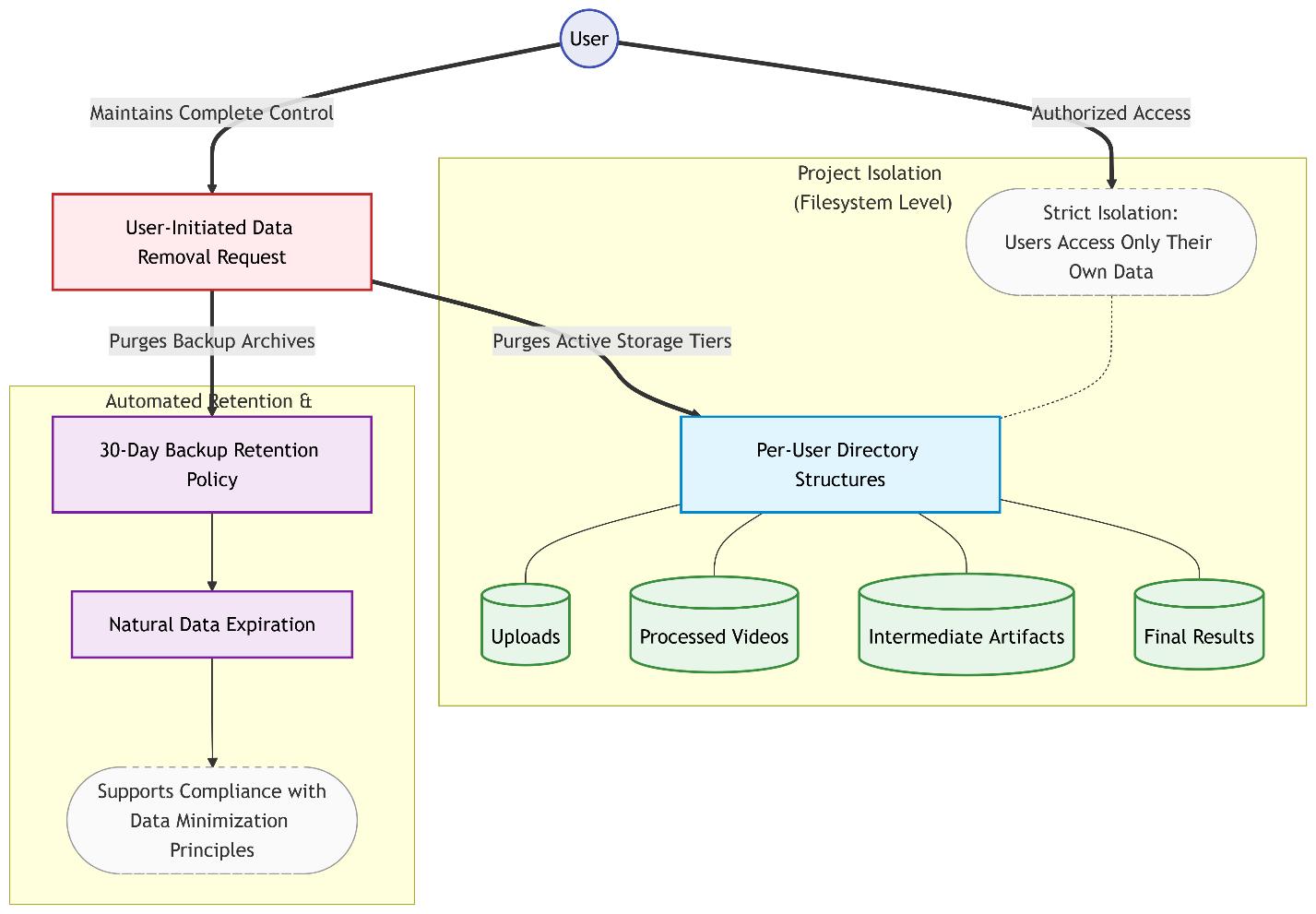

**eMethods 1.4: Network security and access management**

The application implements defense-in-depth network security across multiple layers. External traffic flows through Cloudflare with bot protection, DDoS mitigation, and Web Application Firewall rules before reaching the internal network. Nginx serves as the primary reverse proxy, handling SSL/TLS termination with automated certificate renewal, enforcing HTTPS-only connections, and implementing rate limiting to prevent abuse. The Flask application operates within an isolated Docker network with no direct external exposure; only mounted volumes and explicitly permitted ports are accessible. Authentication is layered through Zitadel, providing enterprise-grade identity management with SSO integration, OAuth 2.0 / OpenID Connect support, and JWT-based session tokens with short expiration windows and refresh token rotation. Additional security layers include IP-based access controls, request validation at each proxy hop, and comprehensive audit logging of all authentication attempts and API access. The architecture follows zero-trust principles, where every request is validated regardless of its origin.

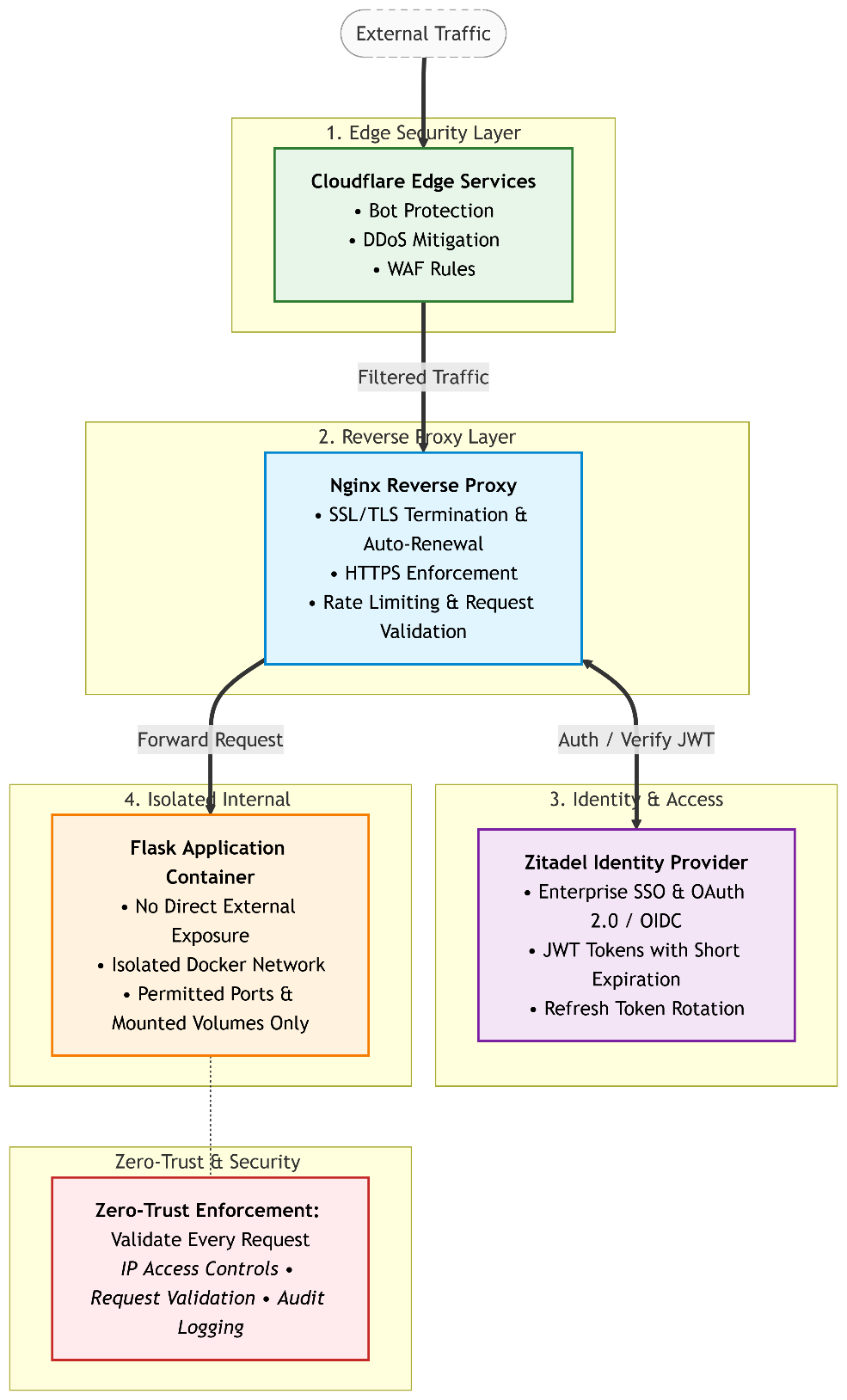

**eMethods 1.5: System performance, latency, and resolution profiling**

To evaluate the real-world operational feasibility of DxTonia Web, we profiled the complete processing pipeline across multiple video resolutions and framerates. Specifically, we benchmarked the standard clinical recordings from Center 1 (Panasonic AG-AC160AP, 1920x1080, 30fps, n=193), smartphone-based recordings from Center 2 (Google Pixel 5, 3840x2160, 60fps, n=105) and down sampled videos from Center 2 (1280x720, 30fps, n=105). Testing was performed on a server equipped with an Intel Xeon Gold CPU and an NVIDIA RTX 4090 GPU (24GB VRAM). The overall latency, as well as the discrete processing times for metadata correction (FFmpeg), keypoint extraction (OpenPose), temporal tracking and patient detection, and neural network inference (dystonia detection ensemble model and DxToniaNet), are reported in Supplementary eTable 2.

**eMethods 1.6: Effect of down scaling on dystonia estimation performance**

Because OpenPose skeleton extraction scales exponentially with pixel density, high-resolution smartphone videos (4K @ 60fps) present a major computational bottleneck. To investigate whether down sampling could accelerate clinical workflows without compromising diagnostic output, we down sampled native 4K smartphone videos from Center 2 to 720p. We then processed these down sampled tracks through the identical severity estimation pipeline and compared DxToniaNet’s performance against native 4K outputs by calculating Pearson correlation (r) and Mean Absolute Error (MAE) across (Supplementary eTable 3).

**eMethods 1.7: Duration scaling and concurrency load testing**

To verify the pipeline's stability and verify the performance of the Redis background job queue, we measured processing latency across varying video durations (ranging from short 5-second Center 1 clips to long 60-second Center 2 recordings). Furthermore, concurrent user load was evaluated by submitting 1and 20 simultaneous upload-and-analysis requests to the server, measuring the queue queue-wait times and overall computational throughput under heavy load. We found that the Redis queue managed the jobs without any failure. However, since the resources server were limited, the processing time increased as the number of users increased.
